# Genetic determinants of gene expression noise and its role in complex trait variation

**DOI:** 10.1101/2024.11.29.24318180

**Authors:** Yuexuan Long, Xiaolin Ni, Tingwei Chen, Qiyang Hong, Jixin Wang, Cong Wang, Zigeng Huang, Haiqing Xu, Mengyi Sun, Junling Pang, Jiyeon Choi, Tongwu Zhang, Erping Long

## Abstract

Even genetically identical cells in a homogeneous environment can exhibit heterogeneous mRNA abundance because of widely unavoidable random fluctuations, typically referred to as ‘gene expression noise’. Recent studies showed that noise, not just a nuisance, is indeed involved in cellular activities (e.g., immune response), evolutionary processes, and diseases mechanisms. However, determinants of the gene expression noise and its functional role in variations of human complex traits remain largely unexplored. Here, we established an atlas of gene expression noise from 1.23 million human peripheral blood cells of 981 individuals, identifying its age- and gender-dependent pattern. We then identified 10,770 independent expression noise quantitative trait loci (enQTLs) for 6,743 unique enGenes (genetically driven gene expression noise) across 7 immune cell types. Most enQTLs were distinct from expression quantitative trait loci (eQTLs) and showed differential enrichment of functional elements across the genome. Colocalization of enQTLs with trait-associated genetic loci interpreted previously unexplained loci and pinpointed novel putative genes underlying hematopoietic traits and autoimmune diseases. Overall, this study unravels the genetic determinants of gene expression noise and implicates as a previously underappreciated mechanism underlying variation of human complex traits and diseases.

## INTRODUCTION

The intrinsic stochasticity of biochemical reactions contributes to a wide distribution of gene expression across a seemingly homogeneous population of cells. This phenomenon of transcriptional variability, called ‘gene expression noise’, has been widely observed in prokaryotic and eukaryotic systems (*1*). Classically, noise can be quantified by time-resolved reporter assay of individual genes (*2*). More recently, single-cell techniques, along with statistical approaches, have been applied to distinguish true biological cell-to-cell variability from measurement errors (*3*). Fueled with these methodological advances, several genetic and epigenetic features were identified to be the potentially deterministic components of noise. In budding yeast, less-active genes with high nucleosome occupancy close to the transcriptional start site (TSS) and genes with presence of TATA box in their promoter sequences are associated with elevated noise (*4*, *5*). In mammalian cells, the association between TATA box and noise has been consistently reported (*6*). Moreover, genes with the absence and shorter length of promoter-region CpG islands were associated with attenuated noise (*7*, *8*). However, our understanding of the interindividual determinants (e.g., the genetic variants across human genome) of noise remains unknown.

Gene expression noise is suggested to be involved in a variety of important biological and pathological processes. For example, noise is elevated in early pluripotent cells and attenuated in later embryonic development stages (*9–11*). In terms of senescence, transcriptional variability increases with age and the disruption of noise control may lead to abnormal immune responses (*12*). Moreover, cell cycle and gene expression noise were intimately connected, as the genes regulating cell cycle were variably expressed across different phases but stable within the same phase (*13*). Most human phenotypes and diseases are complex traits that are contributed by genetic variants from numerous loci across genome (*14*) with mediators of molecular traits (*15*). However, current studies of molecular quantitative trait loci (QTL) mainly focused on steady-state average abundances of a trait (e.g., gene expression); whether and to what extent the variability of a trait (e.g., noise) may contribute to human complex traits remains to be explored.

To address the above fundamental questions, we leveraged the population-scale single-cell datasets of human peripheral blood cells to characterize the transcriptome-wide noise atlas. We then identified 10,770 independent expression noise QTLs (enQTLs) across 7 immune cell types, which were largely independent from expression QTLs (eQTLs) and can explain a substantial proportion of previously unexplained immune trait-associated loci from GWAS. Overall, this study demonstrated the genetic architecture of gene expression noise and highlighted the important role of noise underlying genetic associations with complex traits.

## RESULTS

### Gene expression noise across cell types and associations with age and gender

We curated a single-cell transcriptome atlas from the OneK1K cohort, comprising 1,233,834 peripheral blood mononuclear cells (PBMCs) from 981 healthy individuals of Northern European ancestry (*16*). To accurately calculate the gene expression noise by using a sufficiently high cell number per individual, we grouped the cells into eight major cell types based on established annotations: CD4^+^ T (CD4) cells, CD8^+^ T (CD8) cells, natural killer (NK) cells, B cells, monocytes (Mono), gamma-delta T cells (gdT), dendritic cells (DC), and plasmablasts (plasma) (**Fig. S1A**, **Table S1**)(*16*). The numbers of cells in each cell type ranged from 3,754 (0.3% of the total cells) for plasma cells to 624,570 (50.6%) for CD4 cells (**Fig. S1A**). The dimensionality reduction of Uniform Manifold Approximation and Projection (UMAP) and clustering analysis revealed a clear hierarchical architecture among T (CD4, CD8, and gdT), NK, B, DC, Mono and plasma cells (**Fig. S1B**). Additionally, we observed cell-type specific expression of canonical markers for all cell types, reassuring the validity of cell-type annotation (**Fig. S1C**). On average, 1,255 cells were included per individual and 71.7% (703/981) of individuals contain all eight cell types across the cohort (**Fig. S1D, Fig. S1E, Table S2**).

Technical noise is dependent on the average read count of a gene (i.e., mean-variability dependence) in single-cell data (*17*). To deconvolute the biological cell-to-cell noise from technical noise we applied a statistical approach addressing mean-variability dependence of over-dispersed distribution. Namely, we fit the mean and squared coefficient of variation (CV², excluding low-expressed genes with extremely high CV^2^) based on gamma distribution for each individual and calculated the normalized gene expression noise based on residual and logarithmic transformation for each cell type (**Fig. 1A**, **Methods**) (*3*). We observed the significantly positive correlation between cell numbers and fitting performance (*Corr.* = 0.84, *P* ≤ 2.2 × 10^-26^) and retained the individuals with robust fitting performance (adjusted R² > 0.6, **Fig. S2A**. The plasma cells were thus excluded from subsequent analyses because of the limited sample size for further QTL analysis (only 55 individuals with adjusted *R*^2^ < 0.6; the rest of the cell types have 242-981 qualifying individuals) (*18*). The calculated noise exhibited significantly reduced correlation to mean expression in each cell type compared to CV^2^, validating the efficiency of deconvoluting the mean-variability dependence (**Fig. 1B**).

**Figure 1.**
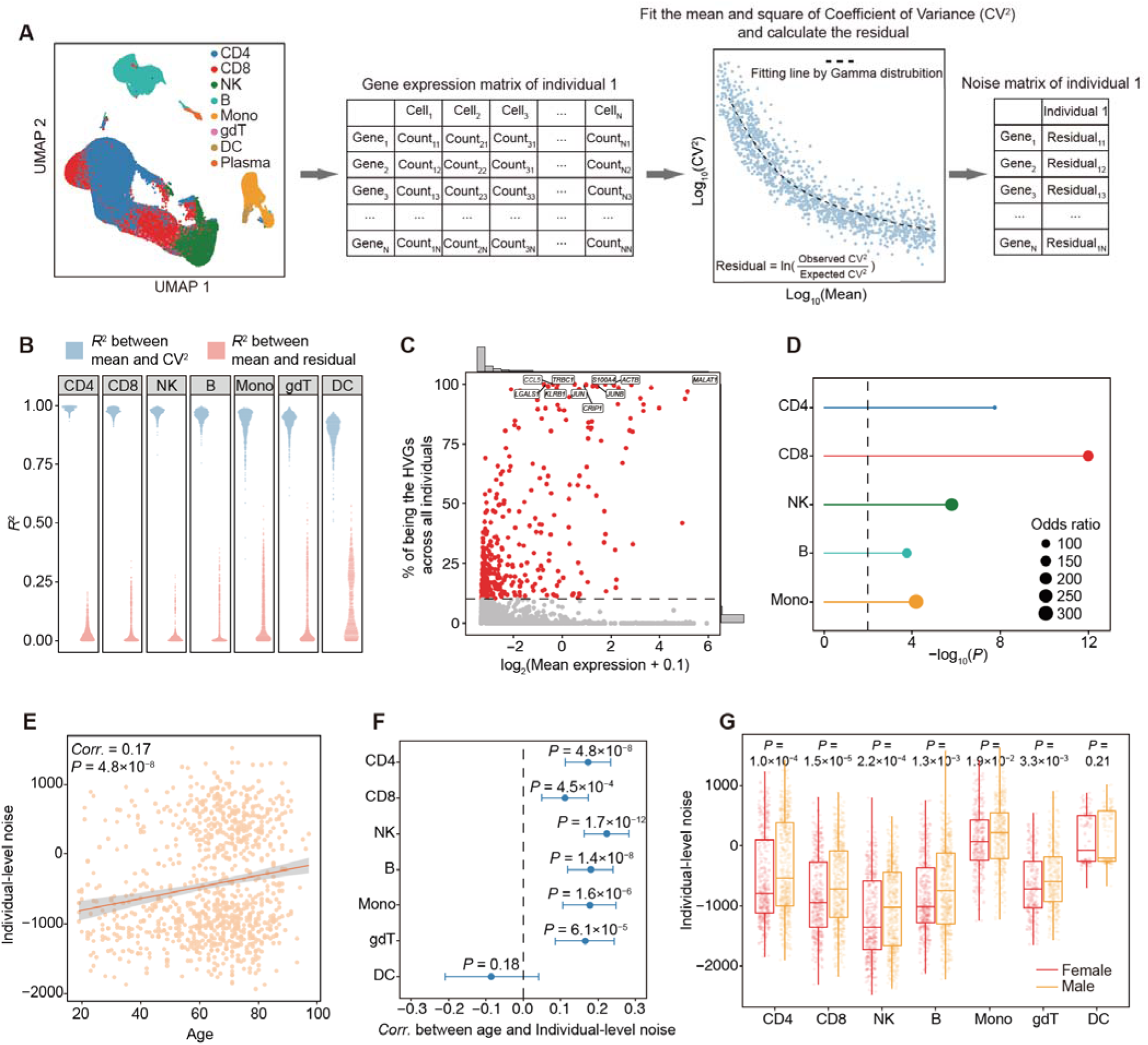
Characterization of gene expression noise, HVGs, and individual-level noise. **(A)** Schematic overview of the framework used to calculate gene expression noise across cell types. The matrixes left and right represent the gene expression and gene expression noise in one cell type of an individual, respectively. The dot plot on the right is the fitted graph of CV^2^ and the mean in one cell type of an individual. A blue dot represents a gene, and the dotted black line represents the Gamma distribution fitting. **(B)** The square of correlation coefficients between mean expression levels and CV^2^ (blue) or residual (red) were calculated using Pearson’s product-moment correlation. Each dot represents an individual. **(C)** Distribution of rates of being the HVGs across all individuals for all genes in CD4 cells. The gray line marks the 10% threshold for identification of highly variable genes (HVGs). Red dots represent HVGs, while gray dots indicate non-HVGs. The labeled genes are the top ten genes with the highest rates of being HVGs across all individuals. The bar graph shows the distribution of mean expression (X-axis) and highly expression noise rates (Y-axis). **(D)** The enrichment of lineage-specific genes in HVGs across five cell types with known lineage-specific gene information. The enrichment *P* value was determined by Fisher’s exact test. **(E)** Correlation between age and the sum of residuals in CD4 cells, with *P*-value and correlation coefficient calculated by the Pearson’s product-moment correlation. Each dot represents an individual. The gray shade indicates the 95% confidence interval. **(F)** Pearson correlation coefficients with 95% confidence intervals for the relationship between age and sum of residuals across 7 cell types, CD4 (N = 981), CD8 (N = 981), NK (N = 978), B (N = 973), Mono (N = 716), gdT (N = 580), and DC (N = 242) cells. **(G)** Comparison of the individual-level noise between males and females across 7 cell types, CD4 (N_male_ = 565, N_female_ = 416), CD8 (N_male_ = 565, N_female_ = 416), NK (N_male_ = 562, N_female_ = 416), B (N_male_ = 565, N_female_ = 408), Mono (N_male_ = 397, N_female_ = 316), gdT (N_male_ = 342, N_female_ = 238), and DC (N_male_ = 129, N_female_ = 113) cells. *P*-values are calculated by the two-side Wilcoxon signed-rank test.

We then identified the highly variable genes (HVGs) as the genes exhibiting significantly elevated expression noise (FDR ≤ 0.05) in at least 10% of individuals in each cell type (**Fig. S2B**, **Methods**). A total of 437 HVGs were identified across all cell types, ranging from 6 HVGs in DC cells to 350 HVGs in CD4 cells (**Fig. 1C**, **Fig. S2C, Table S3**). A substantial proportion of HVGs (41.2%, 180/437) are shared in at least two cell types, while the remaining (59.8%, 257/437) are cell-type-specific (**Fig. S2C; Table S3**). Notably, we identified two genes that were HVGs across all cell types, *GNLY* (relevant to innate immune system) (*19*) and *CRIP1* (relevant to cell proliferation and growth) (*20*, *21*) (**Fig. S2C**). As expected, lineage-specific genes are significantly enriched (all *P* ≤ 0.01) in HVGs across cell types, including *GZMK* and *IL7R* in CD4 cells and *XCL1* in NK cells, (**Fig. 1D**, **Table S4**). Moreover, we observed the significant enrichment of HVGs in adaptive immune responses pathways for CD4, CD8, NK, B, and Mono cells, and cell-cycle pathways for CD4 and NK cells (**Fig. S2D**). Together, these findings, consistent with previous studies (*13*, *22*), implied the involvement of gene expression noise in immune and cell cycle process.

To assess the potential impact of age, gender and cell type on gene expression noise, we aggregated the noise of all genes into individual level based on normalized gene expression noise for each cell type. Our first observation was that the individual-level noise elevated as age increased and this trend was consistent across six cell types, with the exception of DC cells (**Fig. 1E-F**), supplementing the existing evidence of noise-age relationship (*12*, *23–25*). Our second observation is the significantly elevated noise among males relative to females across six cell types (**Fig. 1G**). Such trend has been consistently identified in subsets of both autosomal and sex chromosome genes (**Fig. S3A-B**). Moreover, we found no significant differences of age between males and females (**Fig. S3C**), and the noise difference between male and female significant persisted after adjusting the age as a covariate (ANOVA, *P* = 2.36 × 10^-11^), which confirms that the elevated noise among males is genuine. Interestingly, we noted that monocytes exhibited the highest individual-level noise among cell types (**Fig. 1G**), and monocytes are known with its heterogeneity and plasticity in response to environmental dynamics (*26*).

### Identification of expression noise quantitative trait locus (enQTL)

To investigate how genetic variation influences gene expression noise in a cell-type specific manner, we developed a computational pipeline to identify the expression noise QTLs (enQTLs) and their target genes (enGenes) with genetically driven gene expression noise (**Methods**). We identified enQTL using tensor-QTL based on Pearson product-moment correlation and an adaptive permutation scheme (**Methods**) (*27*). Within each cell type, we incorporated the following covariates: sex, age, six genotype-based principal components, and a Bayesian factor (PEER, separately for each cell type, **Fig. S4A**) (*28*) to eliminate unwanted global variation that could obscure the subtle effects of *cis*-acting genetic SNPs.

To determine the significance threshold of enQTL, we examined the distribution of effect sizes and nominal *P* of lead SNPs after permutation, under the assumption that random SNP selection results 50% (null hypothesis) of consistent allelic direction across cell types (*29*, *30*). We selected a *P* threshold of 0.001, where the allelic direction consistency of SNPs most significantly deviated from null hypothesis (*P* = 3.5 × 10^-58^, binomial test) with 89.7% of the SNPs showing a consistent direction (**Fig. 2A**). A total of 10,770 independent enQTLs involving 6,743 unique enGenes across 7 cell types (**Fig. 2B**) were identified, with breakdowns as follows: 2,016 enQTLs and 1,520 enGenes in CD4 cells, 2,497 enQTLs and 2,011 enGenes in CD8 cells, 969 enQTLs and 772 enGenes in NK cells, 2,182 enQTLs and 1,803 enGenes in B cells, 2,304 enQTLs and 2,039 enGenes in Mono cells, 245 enQTLs and 216 enGenes in gdT cells, and 557 enQTLs and 554 enGenes in DCs. Notably, 120 enGenes were identified as HVGs, including immune function genes (e.g., *KRT1* and *GNLY*) (*31*, *32*) and cell cycle genes (e.g., *S100B*) (*33*) (**Fig. 2D-E, Table S5-6**). We performed conditional enQTL identification for five rounds, where lead SNP-gene pairs from earlier rounds were included as covariates in subsequent rounds (**Table S7; Method**). The majority (5,983/6,743, 88.7%) of enGenes had a single independent enQTL while 0.5% (3/554 in DC) to 23.6% (359/1,520 in CD4) of enGenes had multiple independent *cis*-enQTLs, indicating the allelic heterogeneities in these enGenes (**Fig. 2C**, **Fig. S4B**).

**Figure 2.**
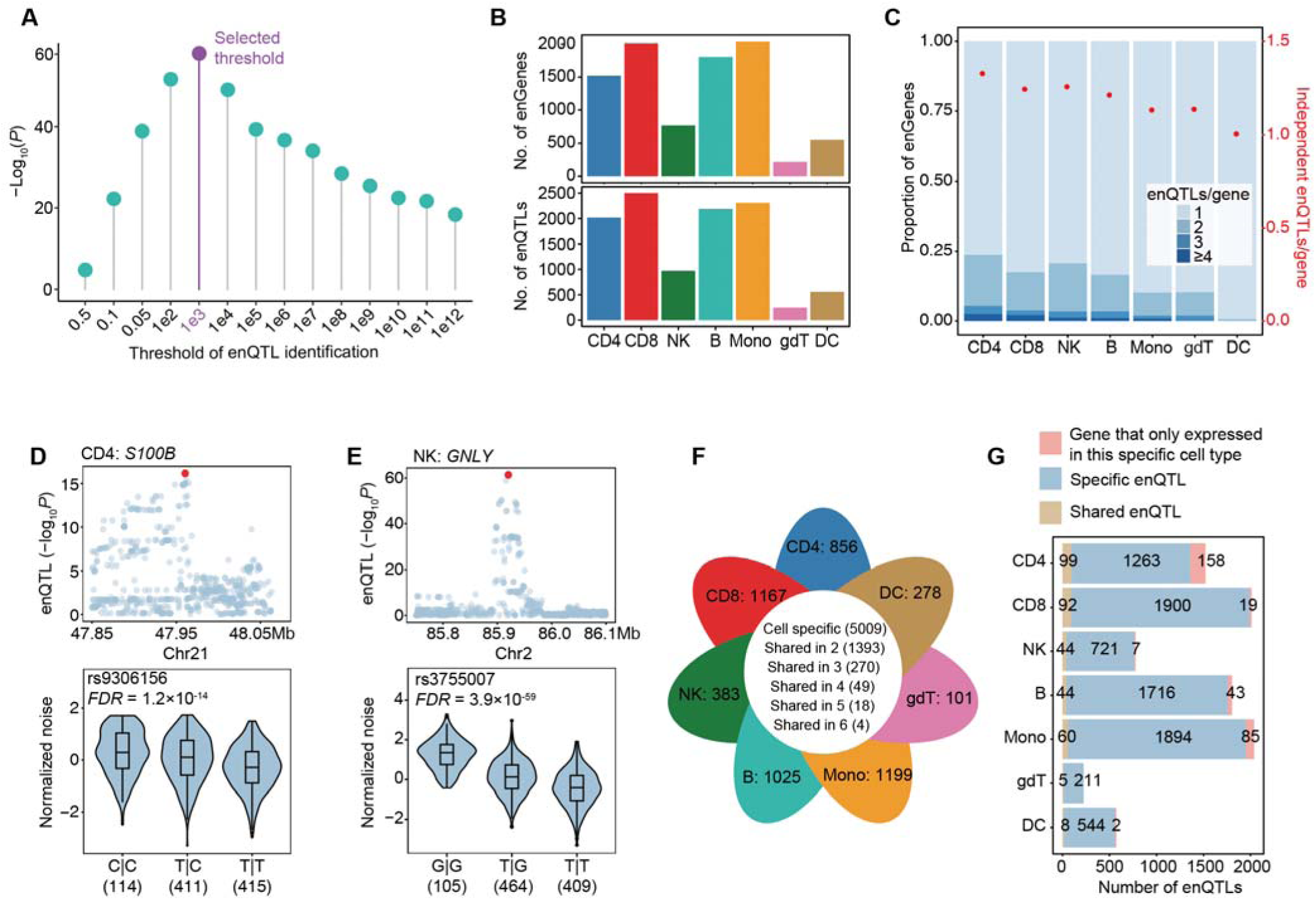
Identification of enQTL across cell types. **(A)** Statistical difference between enQTL directional consistency and null hypothesis (50%) between any of two cell types across a series of thresholds. The threshold of 0.001 was selected (purple). Binomial test was used to determine statistical difference. **(B)** The number of enGenes (top) and enQTLs (bottom) identified across 7 cell types. **(C)** Allelic heterogeneity of enQTLs is depicted by two measures: the proportion of enGenes with one or more independent enQTLs (blue stacked bars; left y-axis) and the mean number of enQTLs per gene (red dots; right y-axis). **(D-E)** Examples of enGenes, including a cell-cycle-related gene *S100B* in CD4 cells **(D)** and an immune-related gene *GNLY* in NK cells **(E)**. In the Manhattan plot, the red dots represent the lead SNPs, with the sample size of each genotype indicated in brackets. **(F)** The number of shared and cell-type-specific enGenes identified across the 7 cell types. The numbers on the petals represent the number of cell-type specific enGenes for each cell type. **(G)** The number of shared and cell-type-specific enQTLs across the 7 cell types. The red segment represents cell-type-specific enQTLs caused by genes only expressed in one cell type.

Of all 6,743 enGenes, 5,009 genes were cell-type-specific enGenes (**Fig. 2F**). Notably, we identified 4 enGenes shared in six cell types, *HLA-C*, *HLA-DQB1*, *EIF5A*, and *RPS26* (*34–37*), which were reported to be involved in immunity and cell cycling (**Fig. S4C**). The lead SNPs targeting the same enGenes in different cell types could be genetically linked. Therefore, we applied the multivariate adaptive shrinkage (MASH) to systematically test the shareability of enQTL across cell types (*38*). We found that 315 enQTLs (3.5%) were shared in at least 2 cell types (**Table S8**). In addition, only a small fraction of the cell-type-specific enQTLs (314, 3.5%) were associated with the enGenes that were specifically expressed in a single cell type (**Fig. 2G**). Together, these results indicated a considerable cell-type specificity in enGenes and enQTLs.

### Independence of enQTL from eQTL

To evaluate the relationship between enQTL and eQTL, we identified the eQTL using the same post-QC data and analytic procedure. We observed that 46.2% (3,122/6,743) of enGenes are eGenes, ranging from 12.3% (68/544 in gdT cells) to 62.4% (949/1,520 in Mono cells) (**Fig. 3A**). Notably, the proportion of overlapping genes showed no significant correlation with the sample sizes in each cell type (*P* = 0.23), suggesting that the overlap is not driven by discovery power. In genes with both an enQTL and an eQTL, most of the lead SNPs (91.8%) for enQTL and eQTL were more than 10 kilobases (kb) away from each other (average distance of 462 kb, **Fig. 3B**) and 89.1% of them were not genetically linked (LD *R*² < 0.2 in 1000G EUR population, **Fig. 3C**). Among the co-lead SNPs (the same lead SNP targeting the same gene), 63.2% (48/76) had consistent allelic directions (e.g., higher average expression with higher expression noise) while 36.8% (28/76) were discordant (**Fig. 3D-E**). We highlighted four different scenarios with specific enQTL (**Fig. 3F**), specific eQTL (**Fig. 3G**), co-lead SNPs with consistent direction (**Fig. 3H**) and discordant direction (**Fig. 3I**). Together, these results highlighted the independence of enQTL from eQTL, implying that *cis*-regulation of gene expression noise is independent from that of mean expression level.

**Figure 3.**
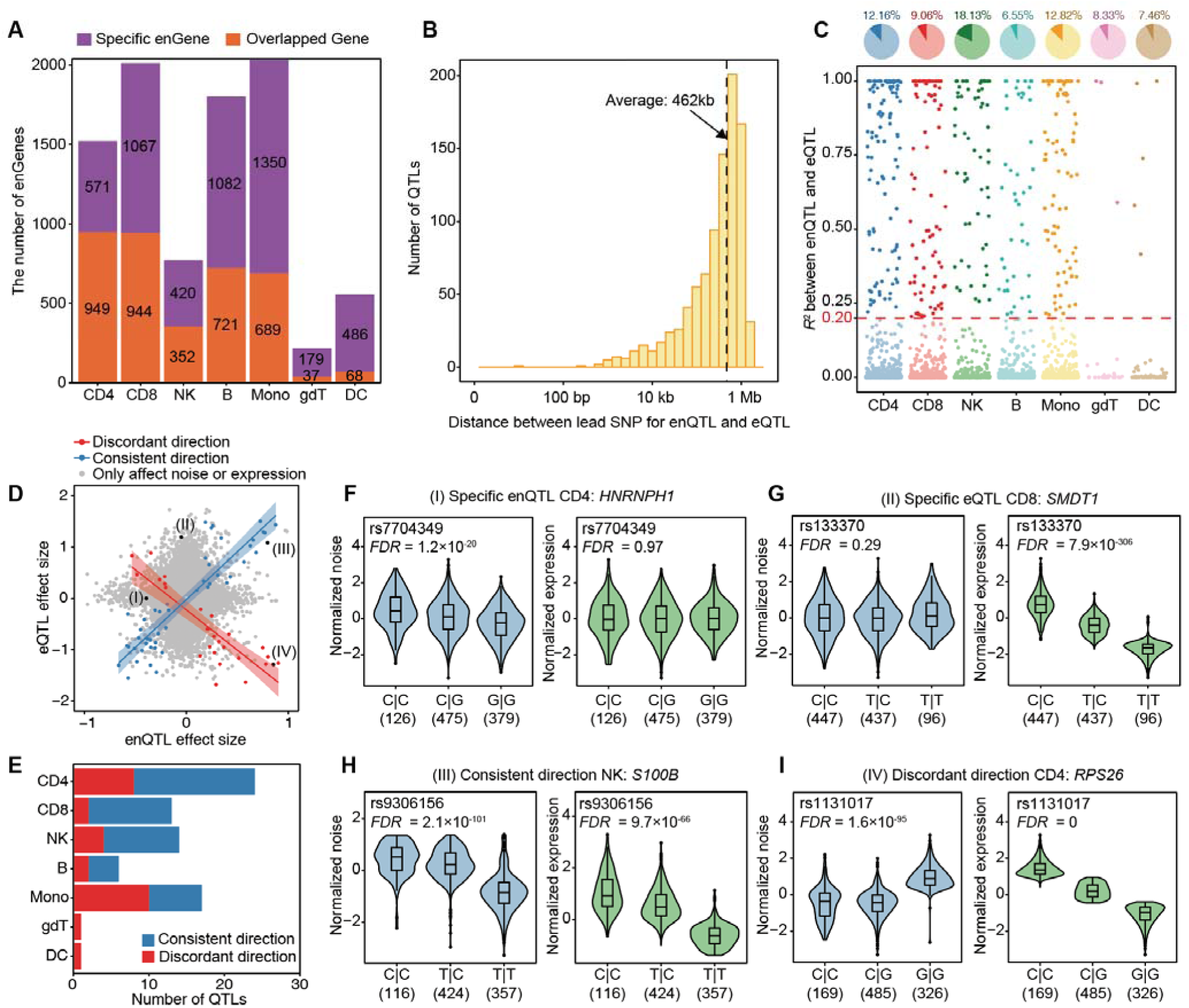
enQTL versus eQTL comparison. **(A)** The number of specific enGenes (purple) and overlapping genes (orange) across the dataset, showing the distribution of genes regulated by enQTLs, eQTLs, or both. **(B)** Distance between lead SNP for enQTL and eQTL of the overlapping genes. The dashed line represents the average distance. **(C)** Linkage disequilibrium (LD) *R*^2^ (1000 Genome, phase 3, EUR population) between the lead enQTL and eQTL SNPs for overlapping genes. The pie chart above indicates the proportion of SNP pairs with *R*^2^ values > 0.2 and ≤ 0.2. Dark and light colors represent SNP pairs > 0.2 and ≤ 0.2, respectively. **(D)** Scatter plot showing the effect sizes of SNPs on gene expression noise (x-axis) and gene expression levels (y-axis), including SNPs affect noise or expression (grey), or both (blue and red colored). **(E)** Number of QTLs affecting both noise and expression with consistent (blue) or discordant (red) directions across cell types. **(F-I)** Four examples were highlighted, including *HNRNPH1* (specific enQTL affecting only gene expression noise), *SMDT1* (specific eQTL affecting only gene expression levels), *S100B* (QTLs with consistent effect direction on noise and expression, and *RPS26* (QTLs with discordant effect directions on noise and expression. The sample size of each genotype was indicated in brackets.

### Functional enrichment and potential mechanisms of enQTL

To investigate the potential mechanisms of enQTLs in noise regulation, we utilized an annotation tool (ANNOVAR), epigenomic and three-dimensional genomic data from the ENCODE project, and enhancer data from SEdb (**Table S9**) (*39–41*). We found that enQTLs are notably enriched in 5’-untranslated regions (5’-UTRs) for all 7 cell types and in exons and upstream/downstream regions of genes for CD4, CD8, and NK (**Fig. 4A**, OR > 1, *P* ≤ 0.01). Additionally, enQTLs were enriched in accessible chromatin regions, active histone modifications (such as H3K27ac, H3K4me3, and H3K4me1), enhancers, and regions associated with chromatin structure, including loop anchors and topologically associating domain (TAD) boundaries for all 7 cell types (**Fig. 4B**, OR > 1, *P* ≤ 0.01). In addition, enQTLs were significantly enriched in regions marked by bivalent modifications in CD4, CD8, B, and Mono (**Fig. 4B**, OR > 1, *P* ≤ 0.05). These results support the involvement of enQTL in transcriptional regulation.

**Figure 4.**
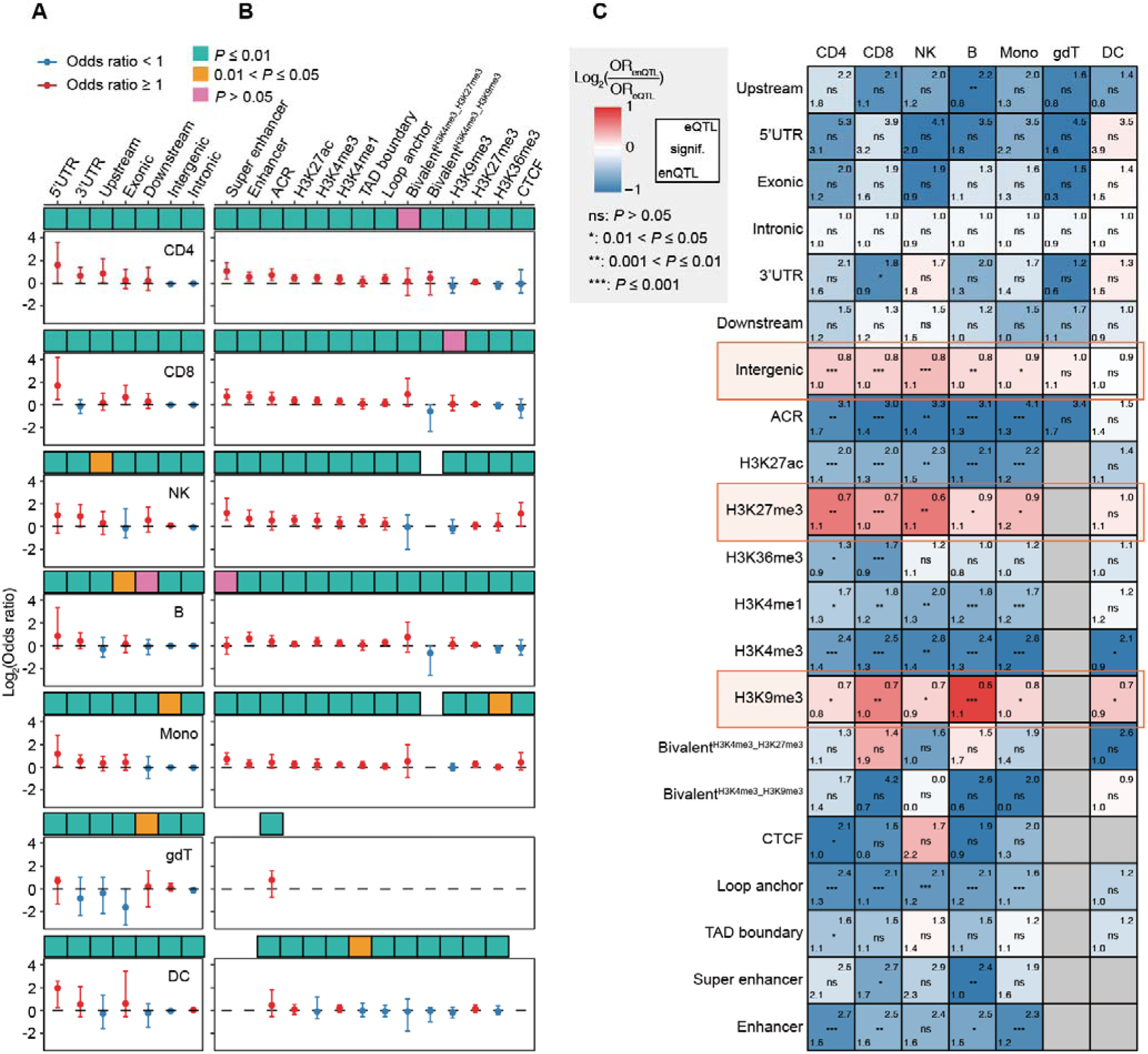
Potential regulatory mechanisms of enQTLs. **(A-B)** Odd ratios (ORs) depicting the enrichment of lead enQTL SNPs in various genomic regions (**A**) and epigenomic regions (**B**) relative to 1,000 permutation sets. Each column represents an estimated OR with error bars indicating the 95% confidence interval. The black dashed line marks the enrichment threshold (OR = 1). The cyan, orange, and pink squares represent *P* ≤ 0.01, 0.01 < *P* ≤ 0.05, and *P* > 0.05, respectively. ORs and *P* values are calculated using Fisher’s exact test. **(C)** ORs of genomic and epigenomic regions comparing enQTLs and eQTLs across cell types. The numbers in the lower left and upper right corners of each square represent the ORs of enQTLs and eQTLs, respectively. The significance in the center of the square indicates the comparison of enQTL and eQTL ORs, with ns, *, **, and *** representing *P* > 0.05, 0.01 < *P* ≤ 0.05, 0.001 < *P* ≤ 0.01, and *P* ≤ 0.001, respectively. The color scale indicates the ratio of ORs between enQTLs and eQTLs: red signifies higher ORs for enQTLs, blue signifies higher ORs for eQTLs, and gray indicates missing annotation data for specific cell type.

Having established that enQTLs are largely distinct from eQTLs, we next explored whether enQTLs and eQTLs display different patterns of functional enrichment. The annotation results from genomic regions revealed that, compared to eQTLs, enQTLs are not significantly depleted in intergenic regions in CD4, CD8, NK, B and Mono (**Fig. 4C**). In terms of epigenetic modifications, eQTLs are significantly depleted in repressive histone modifications of H3K27me3 and H3K9me3 (OR ≤ 1, *P* ≤ 0.05). However, such depletions were not observed for enQTLs, with all ORs ≥ 1 in repressive histone marks (**Fig. 4C**). In general, the enrichment of active regions in enQTLs is attenuated relative to eQTLs (**Fig. 4C**), indicating that repressive chromatin regions and intergenic areas do not adversely affect enQTLs. Together, our data implicated that enQTLs and eQTLs may be mediated by different mechanisms, such as transcription initiation and chromatin remodeling.

### enQTL help interpret GWAS loci

We next asked whether the enQTLs can help uncover the potential mechanisms for trait-associated GWAS loci. We performed the colocalization analysis across 29 hematopoietic traits and 5 autoimmune diseases, assessing whether the enGene-trait association is driven by the same set of casual SNPs (**Methods**). In total, we identified 214 enQTL-colocalized events (unique enGene-trait-cell-type pairs) for blood traits and/or complex diseases (PP.H4 > 0.7, **Table S13, Fig. S5A**). The same pipeline was applied to identify 1,473 eQTL-colocalized events (**Table S14**). We observed that 93.93% (201/214) of the enQTL-colocalized events are independent from eQTL-colocalized events (**Fig. 5A**).

**Figure 5.**
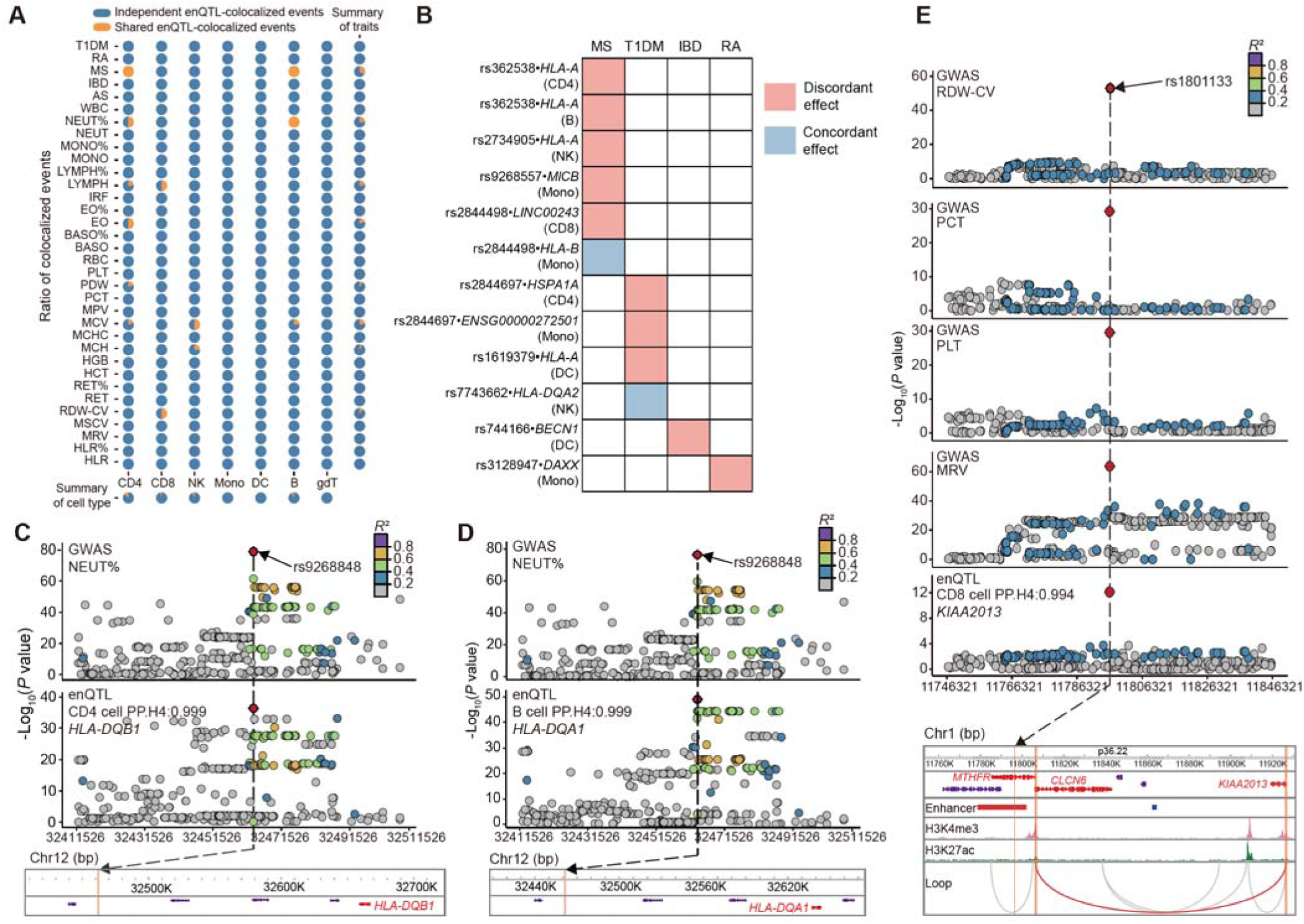
Colocalization between enQTL and GWAS signals of complex traits and diseases. **(A)** The pie chart presents the fraction of enQTL-colocalized events that are independent (blue) or shared (yellow) with eQTL-colocalized events across cell types and traits. **(B)** The heatmap presents the allelic direction between enQTLs and GWAS of 12 autoimmune-disease colocalized events, including discordant (disease-risk allele to attenuated noise, red) and concordant effects (disease-risk allele to elevated noise, blue). **(C-D)** The regional association plots of GWAS (NEUT%) and enQTL of the gene *HLA-DQB1* or *HLA-DQA1* within ± 50 kb of rs9268848 in CD4 and B cells, respectively. **(E)** The regional association plots of GWAS (RDW_CV, PCT, PLT, MRV), and enQTL of the gene *KIAA2013* within ±50 kb of rs1801133 is presented. Full-mode UCSC genes tracks, as well as ChIP-seq peaks and Hi-C loops of CD8 cell line from ENCODE are displayed in this region. Three orange shades represent the location of rs1801133, the left and right anchors of the Hi-C loops, respectively. **Notes:** Mean reticulocyte volume, MRV; Plateletcrit, PCT; Platelet count, PLT; Red cell distribution width, RDW_CV; neutrophil percentage, NEUT%.

Notably, we observed that, in 10 of 12 autoimmune-disease colocalized events, the risk allele was associated with attenuated noise of the target genes (**Fig. 5B**). For example, the SNP rs362538 was nominated by the colocalized events between the GWAS of multiple sclerosis (MS) and enQTLs of *HLA-A* in CD4 cells (**Fig. S5B**), with its G allele corresponding to increased risk of MS and attenuated noise, highlighting potentially beneficial aspect of expression noise. There are many other enQTL-colocalized events noted in the MHC regions, which is known to orchestrate the immune response to viral antigens through genetic variations in leukocyte antigen (HLA) genes (*42*, *43*). For example, rs9268848 is nominated from enQTL-colocalized events for the noise of *HLA-DQA1* and *HLA-DQB1*, contributing to the phenotypic variation of neutrophil percentage (**Fig. 5C-D**).

There are several SNPs prioritized by colocalization that have been reported to associate with other human traits outside the 33 tested traits. A well-known obesity-associated SNP, rs8887 (*44*), is nominated by enQTL-colocalized events of target gene *TNFAIP8L1* with GWAS of 5 hematopoietic traits but no eQTL-colocalized events (**Fig. S5C, Table S13**). An ankylosing spondylitis (AS)-associated SNP, rs11616188 (*45–48*), was nominated in the colocalization between GWAS traits of mean corpuscular hemoglobin (MCH) and mean corpuscular volume (MCV) and enQTL of *MLF2*, a Myeloid Leukemia Factor 2 implicated in leukemia (**Fig. S5D-E**). Notably, rs140522 is another SNP prioritized from an enQTL-colocalized event between high light scatter reticulocyte count (HLR) and target gene *TYMP* (**Fig. S5F**). Multiple GWAS reported rs140522 to be significantly associated with leukemia (*49–51*). Therefore, the role of *TYMP* as a dysregulated immunity gene in chronic lymphocytic leukemia could be hypothesized (*52*).

There are two cases of missense SNPs that were nominated from enQTL-colocalized events. The first case is rs1801133, a missense variant of *MTHFR* with known associations of reduced enzymatic activity, higher homocysteine levels, and multiple diseases (colorectal cancer, HBV infection, late-onset Alzheimer’s disease, and age-related disorders) (*53–56*). In our data, rs1801133 is nominated from enQTL-colocalized events in CD8 cells targeting *KIAA2013,* and the cell-line-based ChIP-seq and Hi-C data supported the chromatin interaction between the promoters of *MTHFR* and *KIAA2013* (**Fig. 5E**). Another case is the well-studied missense SNP, rs7412 (ApoE epsilon2) associated with various diseases (cardiovascular disease, hypertension, dementia) and human lifespan by regulating lipid metabolism (*57–61*). In our data, rs7412 was nominated from enQTL-colocalized events in CD8 cells of lncRNA (*CTB-129P6.11*), a novel target that have not been previously prioritized (**Table S13**).

Collectively, our results suggested that many enQTLs contribute to hematopoietic traits and autoimmune diseases, independent of eQTLs, thus providing novel cell-type-specific mechanistic hypotheses for these loci, including candidate causal variants, target genes, and cell types of action.

## DISCUSSION

Understanding the genetic basis of complex traits and diseases is a fundamental question in human genetics and precision medicine. GWAS have identified numerous trait-associated loci and QTL mapping has been proved as a promising approach in interpreting their molecular mechanisms. However, the mainstream QTL focus on steady-state average abundance of a trait (e.g., gene expression for eQTL) and a considerable fraction of trait-associated loci lack eQTL support (*62*). Alternatively, enQTL aim to detect loci that affect the variability in gene expression across individuals, which reflects underlying stochasticity or responses to subtle environmental fluctuations that are not captured in mean expression levels. In this study, we utilized population-scale single-cell data to establish the first-to-date enQTLs dataset across immune cell types in human peripheral blood. Our findings supported that most enQTLs are distinct from eQTLs and can explain a substantial proportion of previously unexplained loci, highlighting noise as an important mediator underlying genetic associations with complex traits such as hematopoietic traits and autoimmune diseases.

The transcriptome-wide noise atlas allows for two important observations. First, we observed consistent pattern of elevated noise with age in six cell types. Previous studies have identified increased variability in the expression of immune response genes, attributing to destabilization of the immune activation program in CD4+ T cells (*12*). Similarly, transcriptional variability was reported to increase with age in the human pancreas, along with increased stress signature and atypical hormone expression (*63*). Here, we expanded the age-noise correlation as a consistent observation to multiple immune cell types, supporting the theory that molecular variability can be regarded as a signature of aging and is comparable across individuals as a quantitative trait. Second, we observed consistent pattern of elevated noise among males relative to female, independent of the aging effect. Previous studies reported an estrogen-dependent control of transcriptional burst in a breast cancer cell line (*64*) and cis-regulatory control of transcriptional noise in response to estrogen (*65*), implying the role of hormone in noise regulation. In the evolutionary perspective, natural selection may favor the attenuated noise in female to ensure more consistent cellular function during critical periods (e.g., pregnancy) while males may potentially tolerate more noise in exchange for other fitness advantages (e.g., risk-taking behaviors) (*66*). In any case, it would be interesting to investigate the in-depth mechanisms of gender-specific noise pattern in future studies.

A key outcome of our work is to provide a set of enQTLs that could be contributing to human complex traits and diseases, which provides novel cell-type-specific mechanistic hypotheses for these loci, including candidate causal variants, target genes, and cell types of action. Noise is, intuitively, regarded as an undesired byproduct of biological systems and often conceives detrimental effects. Unexpectedly, our data supported that autoimmune disease risk variants, in many cases, were associated with attenuated noise of nearby target genes. This unique directional effect implies the beneficial aspect of noise and highlights the evolving understanding of its double-edged role in evolutionary consequences. An interesting hypothesis is that the elevated immune response can be attributed to more sporadic transcriptional expression, such as a subpopulation of early-response cells with highly expressed immune-activation genes (*67*). Individuals with early-response cells were thus characterized with elevated noise and showed higher response strength in immune system. It should also be noted that very few of the enQTL-colocalizing genetic loci overlapped with eQTLs, indicating that enQTLs are largely independent. In the classic ‘random telegraph model’, noise can arise from transcriptional bursting, where genes switch between “on” and “off” states (*68*). The frequency of bursts (how often the gene is turned on) often affects the mean expression level, whereas the size and duration of bursts (how much mRNA is produced when the gene is on) contribute more to noise. These two aspects can be regulated independently. For instance, the gene may be frequently activated (high mean expression) but still produce variable amounts of mRNA during each activation event (high noise). In addition, the independence between enQTL and eQTL can be attributed to distinct biological processes and regulatory mechanisms, such as post-transcriptional control (e.g, mRNA stability) (*69*) and epigenetic dynamics (e.g., bivalent promoters) (*7*). It could be anticipated that this enQTL dataset with full summary statistics could be helpful for future studies to identify functional genes and variants for other complex traits.

Collectively, our study provides comprehensive insights into the genetic determinants of gene expression noise in the human genome and a valuable resource for understanding the role of expression noise in human complex traits and diseases susceptibility. Our findings underscore the importance of noise as a biologically meaningful molecular traits, with the potential of discovering novel mechanisms in transcriptional regulation.

## METHODS

### Quality control of the genotype data

The genotype data were downloaded from Gene Expression Omnibus (GSE196830) (*16*). Intensity data files were transferred to PLINK data format using GenomeStudio PLINK Input Report Plug-in (v2.1.4) (*70*). We excluded the SNPs, if they 1) had missing rate > 3%, 2) with minor allele frequency (MAF) < 0.01, 3) deviated from Hardy-Weinberg equilibrium (*P* < 1 × 10^-3^), 4) with ambiguous strand and flipped SNPs with reverse strand genotypes. We excluded the individuals, if they 1) had missing genotype rate > 3%, 2) with abnormal autosomal heterozygosity (± 3 SD deviated from the mean), 3) one of any pair of individuals with estimated relatedness larger than 0.125, 4) with non-European ancestry (defined as being ± 6 SD from the European mean on PC1 and PC2 based on 1000G EUR individuals). The PCs were calculated using SMARTPCA software (v13050). Ultimately, we obtained 495,631 high-quality SNPs on 22 autosomes from 1,034 individuals.

### Imputation of genotyping data

To enable the required formatting of imputation, we generated the bed and frequency file, executed QC script by HRC-1000G-check-bim.pl utility, and generated vcf file using VcfCooker (v1.17.5). The Michigan Imputation Server (*71*) was used for imputation with the Haplotype Reference Consortium panel (HRC r1.1 2016, EUR population) on each autosomal chromosome, with Minimac4 and Eagle v2.4.1. Finally, we obtained 5,295,853 SNPs with imputation quality *R*^2^ > 0.8 and MAF > 0.05 for subsequent analysis.

### Expression data processing

We downloaded the processed data of OneK1K cohort from the Human Cell Atlas (HCA) portal. SCTransform (v0.4.1) (*72*) was used to normalize the gene UMI count matrix, accounting for the confounding effects from sequencing depth, pooling batch, library size, mitochondrial mapping percentage, and other latent factors. Since the calculation accuracy of gene expression noise is highly correlated with the cell number of each cell type, we combined the published cell clustering and annotation into 8 cell types (CD4, CD8, NK, B, Mono, gdT, DC, and plasma) (*16*).

### Gene expression noise calculation

To deconvolute the gene expression noise from the mean-variability dependence in single-cell data, we applied a quantitative statistical approach (*3*) with the following steps.

For the first step. we filtered the individuals if the cell counts of an individual in a specific cell type is less than 3 or the total number of expressed genes is less than 100. For all the included individual of a given cell type, we calculated the average expression and the observed expression square of coefficient of variation (CV^2^) of each gene.

For the second step, we applied the gamma distribution of generalize linear model to fit the mean gene expression and CV^2^ of all genes at individual level for each cell type. We defined a minimum value of gene expression (the 85^th^ percentile among all genes with CV^2^ > 0.3) to exclude low-expressed genes with extremely high CV^2^. We removed genes with abnormal CV^2^ (± 3 SD from the mean) as well. The starting intercept and slope were set as 0 and 0.5, respectively. Gamma fitting is the default parameter except using the identity link. To evaluate the fitting performance, we calculated the adjusted *R*^2^ based on the sum of squares total, the sum of squares error, and the number of genes used for fitting.

The fitting curve of the gamma distribution represented the expected CV^2^ of each gene and we calculated the residual-based noise by 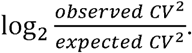 If observed CV^2^ is N/A (possibly caused by zero expression level in all cells), the residual is assigned as a missing value (N/A).

### HVG and individual-level noise calculation

We estimated the statistical significance of deviation of each gene relative to expectation by the chi-square test and the resultant *P* values was adjusted by the Bonferroni correction (FDR). The HVGs were defined as the genes that were significantly deviated (FDR ≤ 0.05, higher than expected) in more than 10% of individuals within a cell type. HVGs were calculated separately for each cell type. The individual-level noise was calculated by aggregating the noises of all genes within an individual.

### enQTL identification

We used tensorQTL (v2.0.0) (*27*) for enQTL identification with covariates of the age and sex, the top six genotype PCs, and latent PEER factors (details in the following section). We restricted our search to SNPs within +/- 1Mb of transcription start site (TSS) of a gene. The nominal *P* value of lead SNP was extrapolated based on Pearson product-moment correlation and an adaptive permutation scheme by tensorQTL (parameter --permute 1,000, --maf_threshold 0.05). We corrected the nominal *P* value with the Benjamini– Hochberg procedure (method = “fdr”). Significance of the resulting SNP-enGene pairs was determined at a false discovery rate (*FDR*) threshold of 0.001 within each chromosome. To explore the conditionally independent signals of enQTLs within each locus, we performed the conditional analysis by regressing out the effect of the lead SNP. Specifically, we obtained the lead SNP of first round enQTL analysis and repeated the QTL mapping to identify the secondary enQTLs by adding the first-round lead SNP as a covariate. Conditional analysis was repeated up to five rounds. We focused on genes that were expressed in at least 10% of the individuals in each cell type. For eQTL, we applied the same procedure to enable the fair comparison. Multivariate adaptive shrinkage (MASH) (*38*) was employed to assess enQTL sharing across cell types. An enQTL was classified as shared between two cell types if the local false sign rate (lsfr) of slopes was ≤ 0.1 in both cell types.

### Latent factor identification and removal

We employed PEER (v1.0.0) (*28*) to identify the latent covariates of unwanted variance stemming from non-genetic factors. Prior to peer factor inference, the expression noise matrices were processed through inverse rank normalization. The K-Nearest Neighbors algorithm was applied to impute the missing values in the matrix of gene expression noise. Distance estimations were used to identify K individuals that are spatially similar or close to each other in the dataset. These K individuals were then used to estimate the missing value and to impute as the average of the K neighborhoods found in the dataset. We determined the number of PEER factors to include based on the number of identified enQTL and correlations among PEER factors (**Fig. S4**), which were the top three, four, three, six, four, one, twenty PEER factors for CD4, CD8, NK, B, Mono, gdT and DC, respectively.

### Functional annotation and enrichment analysis of enQTL

The annotation of enQTLs and eQTLs involves two sections. First, we used the annotate_variation.pl script from ANNOVAR (*39*) to annotate all SNPs after imputation and filtering described above, applying the default annotation databases, except for -geneanno and - buildver hg19. Second, we collected publicly available epigenomic data for each immune cell line, mainly from two databases: the ENCODE Portal (*40*) and the Super-Enhancer database (SEdb) (*41*). From the ENCODE project, we downloaded pseudo-replicated peaks files (in narrowPeak format) of ATAC-seq and ChIP-seq (CTCF, H3K4me3, H3K9me3, H3K27me3, H3K36me3, H3K4me1, H3K27ac) and TAD and Loop files (in bedpe format) of intact Hi-C. From SEdb, we downloaded super-enhancer and enhancer files (in bed format). It is worth emphasizing that the data from ENCODE were based on the hg38 reference genome, so we used liftOver to convert them to the hg19 reference genome. The TAD boundary region is defined as the region extending 10 kb (two resolutions) on either side of the TAD boundary. Loops with an fdrBL ≤ 0.01 are considered significant loops, and ChIP-seq peaks with a Q-value less than 0.01 are defined as significant peaks.

The enrichment analysis of enQTL was performed in the following steps. First, we annotated lead SNPs of enQTL, respectively in different functional annotation categories (e.g., H3K4me3 peaks) based on their physical positions and quantified the proportion of enQTL SNPs in each category. Second, to ameliorate ascertainment bias, we randomly sampled at the same number of lead SNPs of enQTL (a single SNP per gene within +/- 1Mb of TSS) and quantified the proportion of enQTL SNPs in each category as the negative control. This sampling procedure was repeated 1,000 times. Third, we applied the Fisher’s exact test in each functional annotation category as the ratio of the proportion of enQTL SNPs in a functional category over the mean proportion of the control SNPs in the category across 1,000 replicates, resulting in final *P*-values and OR. The significant enrichment was defined as OR > 1 and *P* ≤ 0.05. The same Fisher’s exact test was used for the enrichment comparison between enQTLs and eQTLs.

### Colocalization analysis between enQTL and GWAS loci

Colocalization analysis between enQTLs and GWAS signals was carried out using a Bayesian framework for colocalization (coloc) (*73*). A total of 29 hematopoietic traits and 5 autoimmune diseases were involved (full names and abbreviations in **Table S10**). First, we selected the GWAS loci at genome-wide significance of *P* < 5 × 10^-8^ and clumped by LD *R*^2^ < 0.1 and region size of 100kb. Second, we selected the significant enQTL at *FDR* < 0.001 and overlapped with GWAS loci. Third, we performed colocalization for all enQTL-GWAS-overlapped loci. The colocalization statistics PP.H4 (posterior probability of both traits being associated with the same causal SNP) was calculated within the region of 100kb (± 50 kb) of the GWAS lead SNP. PP.H4 > 0.7 were defined as the GWAS-QTL colocalized events. The colocalization of eQTL and GWAS was performed using the same procedure.

## DATA AVAILABILITY

The following publicly available datasets were used in this study: GWAS summary statistics of blood traits, ftp://ftp.sanger.ac.uk/pub/project/humgen/summary_statistics/UKBB_blood_cell_traits; IBD, https://ftp.sanger.ac.uk/pub/project/humgen/summary_statistics/human/2016-11-07; SLE, http://urr.cat/data/GWAS_SLE_summaryStats.zip; MS, http://imsgc.net/data/imsgc_mssev_discovery.tar.gz. The GWAS Catalog accession numbers for summary statistics: RA, GCST90132001-GCST90133000; AS, GCST005001-GCST006000; T1DM, GCST90014001-GCST90015000; AD, GCST90012001-GCST90013000. The single-cell RNA-seq data of the OneK1K cohort, https://cellxgene.cziscience.com/collections/dde06e0f-ab3b-46be-96a2-a8082383c4a1. OpenTarget Genetics, https://genetics.opentargets.org. Additional details of the data used in this work are provided in the paper and supplementary data.

## SOFTWARE AND CODE AVAILABILITY

We performed our analyses using the following publicly available software packages: LDlink, https://ldlink.nci.nih.gov/; UCSC liftOver, http://genome.ucsc.edu/cgi-bin/hgLiftOver; PLINK v2.0: https://www.cog-genomics.org/plink/2.0; coloc v5.2.3, https://github.com/chr1swallace/coloc; ANNOVAR, https://annovar.openbioinformatics.org/en/latest; SMARTPCA v13050, https://github.com/chrchang/eigensoft; VcfCooker v1.17.5, https://genome.sph.umich.edu/wiki/VcfCooker; PEER: https://github.com/PMBio/peer, tensorQTL v2.0.0, https://github.com/broadinstitute/tensorqtl; Snakemake v5.4.0, https://github.com/snakemake/snakemake; SCTransform v0.4.1, https://github.com/satijalab/sctransform.

## Acknowledgement

E. Long is supported by the National Natural Science Foundation of China (Excellent Youth Scholars Program, 32300483, 32470635, 82090011), State Key Laboratory Special Fund (2060204), and Chinese Academy of Medical Sciences Innovation Fund (2023-I2M-3-010, 2023-I2M-2-001). We thank Xushen Xiong, Jianzhi Zhang, Zhiyue Zhang, Choi lab members, and Long lab members for valuable comments. We thank the Bioinformatics Center of Institute of Basic Medical Sciences for computing support and Furen Shi for technical assistance.

## Author contributions

E. Long, Y. Long, and T. Zhang contributed to the conception and design of the work. Y. Long, X. Ni, T. Chen, Q. Hong, J. Wang, and C. Wang contributed to the data acquisition, curation, and analysis. Z. Huang, H. Xu, M. Sun, J. Pang, and J. Choi provided valuable assistance. All authors contributed to the drafting and revising of the manuscript.

## SUPPLEMENTARY FIGURES

**Figure S1.**
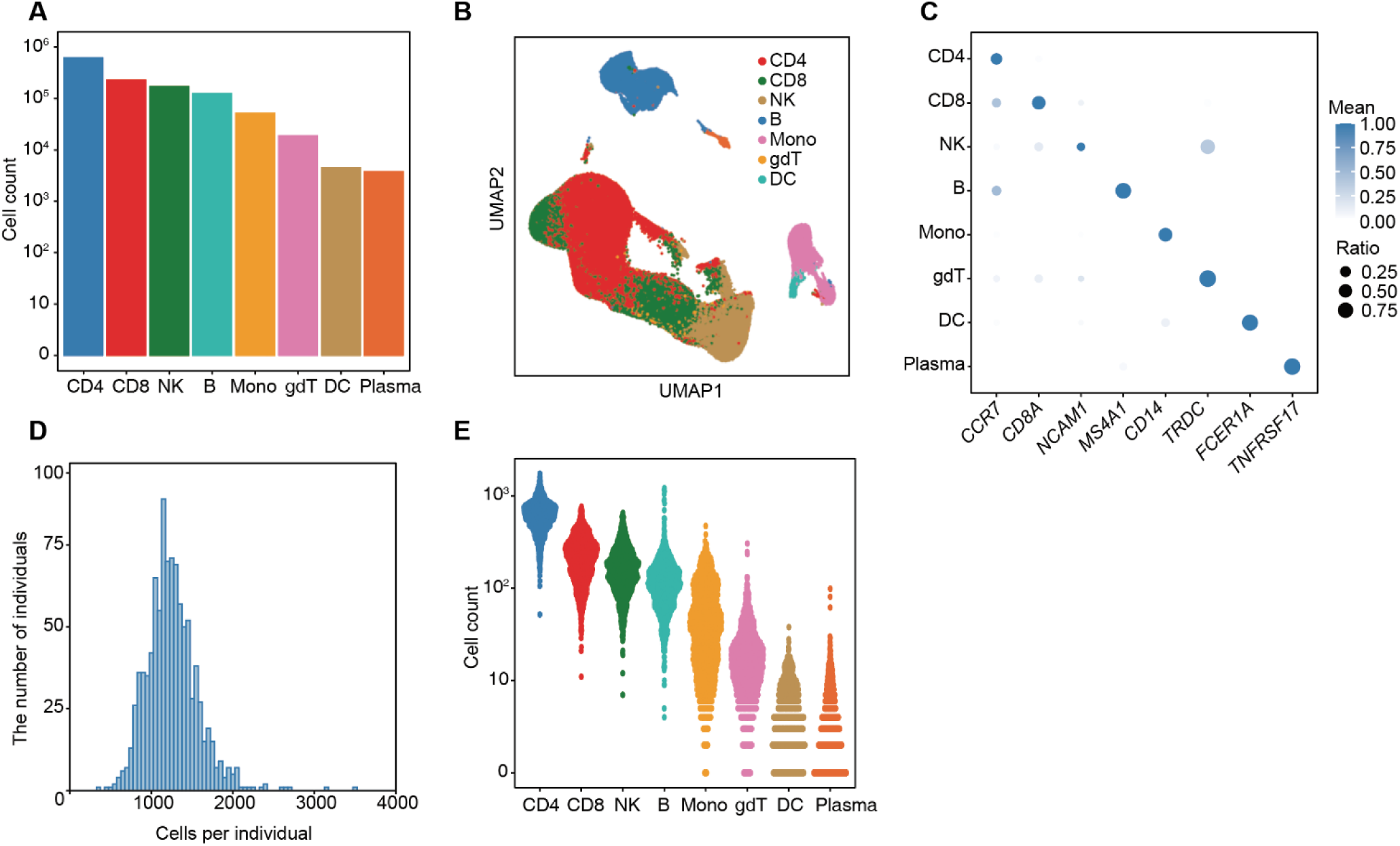
Classification and composition analysis of cell types. **(A)** The total number of cells classified into each cell type. **(B)** UMAP plot displaying single cells from all individuals, highlighting eight transcriptionally distinct populations. **(C)** Dot plots for differentially expressed canonical markers of peripheral immune cells, showing strong concordance with known canonical markers. The color represents the mean level of gene expression. The dot size represents the ratio of individuals with expressed gene relative to total number of individuals. **(D)** Average number of individual cells per donor, ranging from 330 to 3,480 cells. **(E)** The distribution of total number of cells per individual for each cell type.

**Figure S2.**
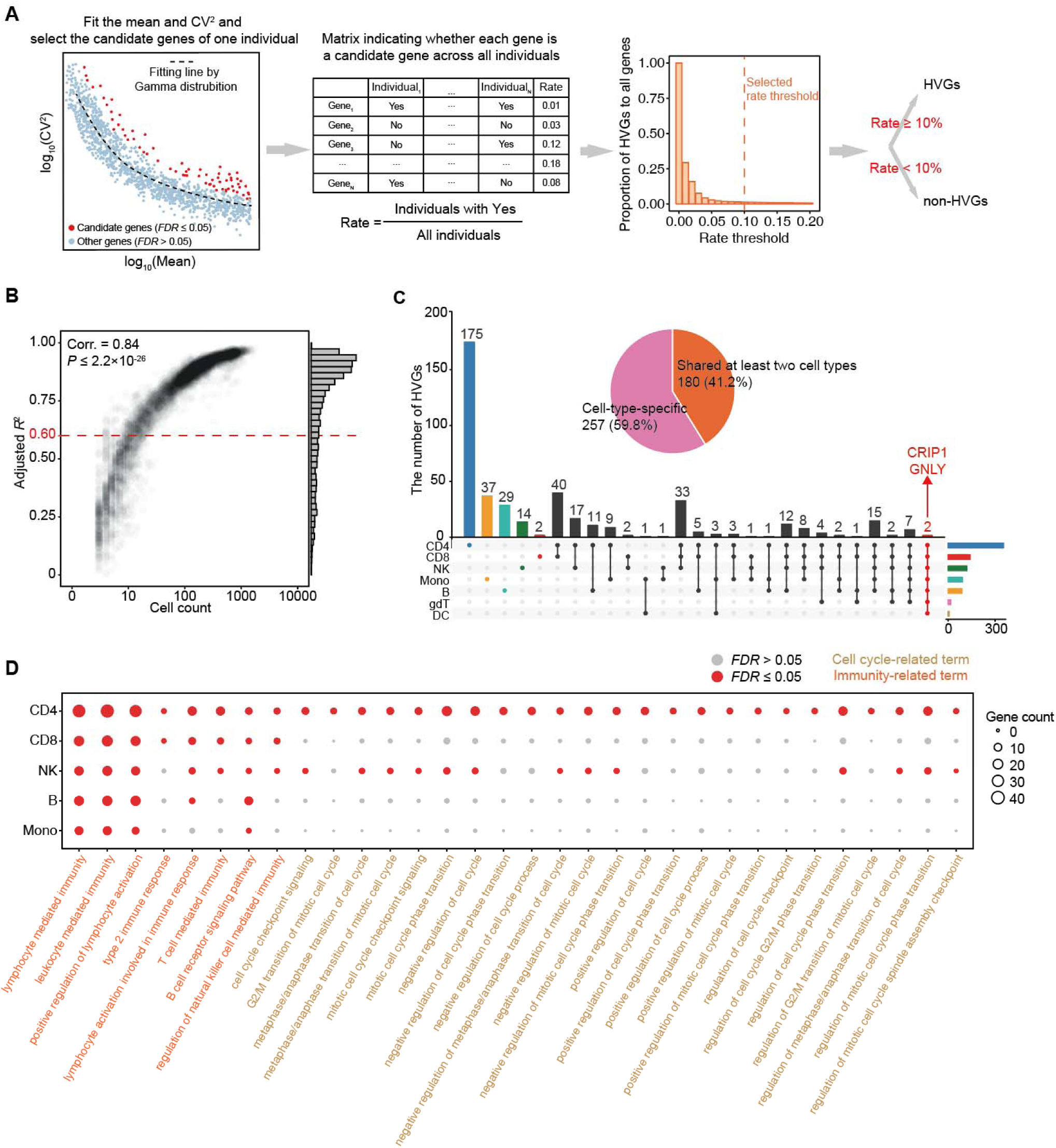
Identification and functional enrichment of HVGs. **(A)** The workflow for HVG identification. The left-side dot plot represents the fitted graph of CV^2^ and the mean and selection of the candidate genes in a single cell type of an individual. Blue and red dots represent candidate genes (*FDR* ≤ 0.05) and other genes (*FDR* > 0.05), respectively, and the dotted black line represents the result of the Gamma distribution fitting. The matrix represents whether each gene is a candidate gene across all individuals within one cell type. The bar plot represents proportion of HVGs to all genes across a series of rate thresholds. HVGs in this cell type were defined as being HVGs in more than 10% (selected rate threshold) of individuals. **(B)** The correlation between cell counts (>2) and adjusted *R*^2^ (fitting effect) across cell types. The correlation coefficient and *P* value are calculated using Pearson’s product-moment correlation. **(C)** The distribution of HVGs across cell types. Lines connect HVGs identified in multiple cell types. The x-axis represents all combinations, and the total number of HVGs detected per cell type is shown on the right side. The red-highlighted dots represent 2 genes shared across all cell types. The pie chart illustrates the number of shared (orange) versus cell-type-specific (pink) HVGs. **(D)** GO enrichment results for HVGs across 5 cell types with number of HVGs > 20. Dot size represents the number of genes in each GO term. Red and grey dots indicate significantly and non-significantly enriched GO terms (*FDR* ≤ 0.05), respectively. Orange and brown labels represent pathways related to immunity and the cell cycle, respectively.

**Figure S3.**
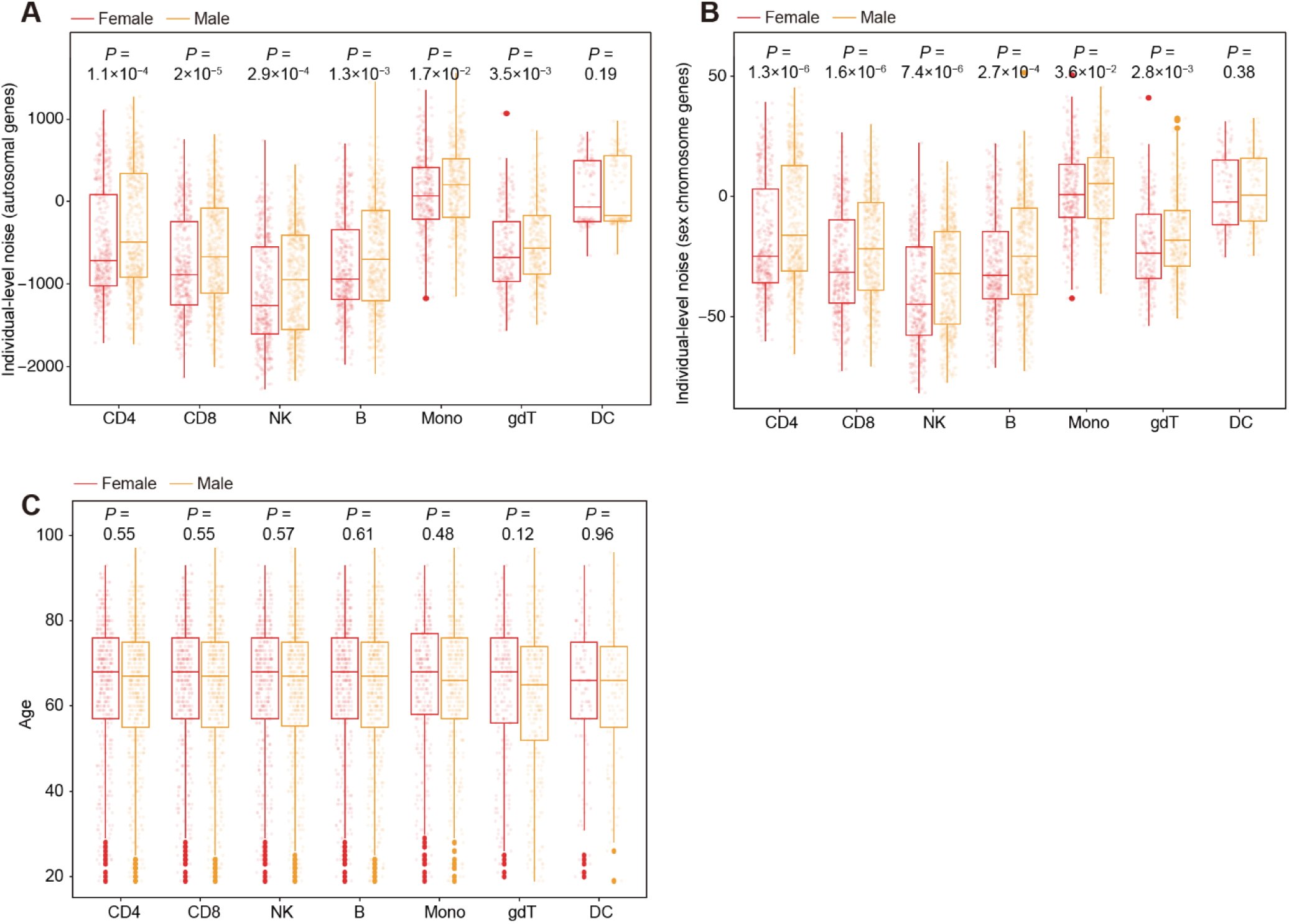
Correlation between age, sex, and gene expression noise. **(A-B)** Comparison of residual sums between males (yellow) and females (red) for autosomes **(A)** and allosomes (including both chrX and chrY) **(B)**. Statistical significance was calculated by the Wilcoxon signed-rank test. **(C)** The age distribution between males and females with *P*-value, also assessed using the Wilcoxon signed-rank test.

**Figure S4.**
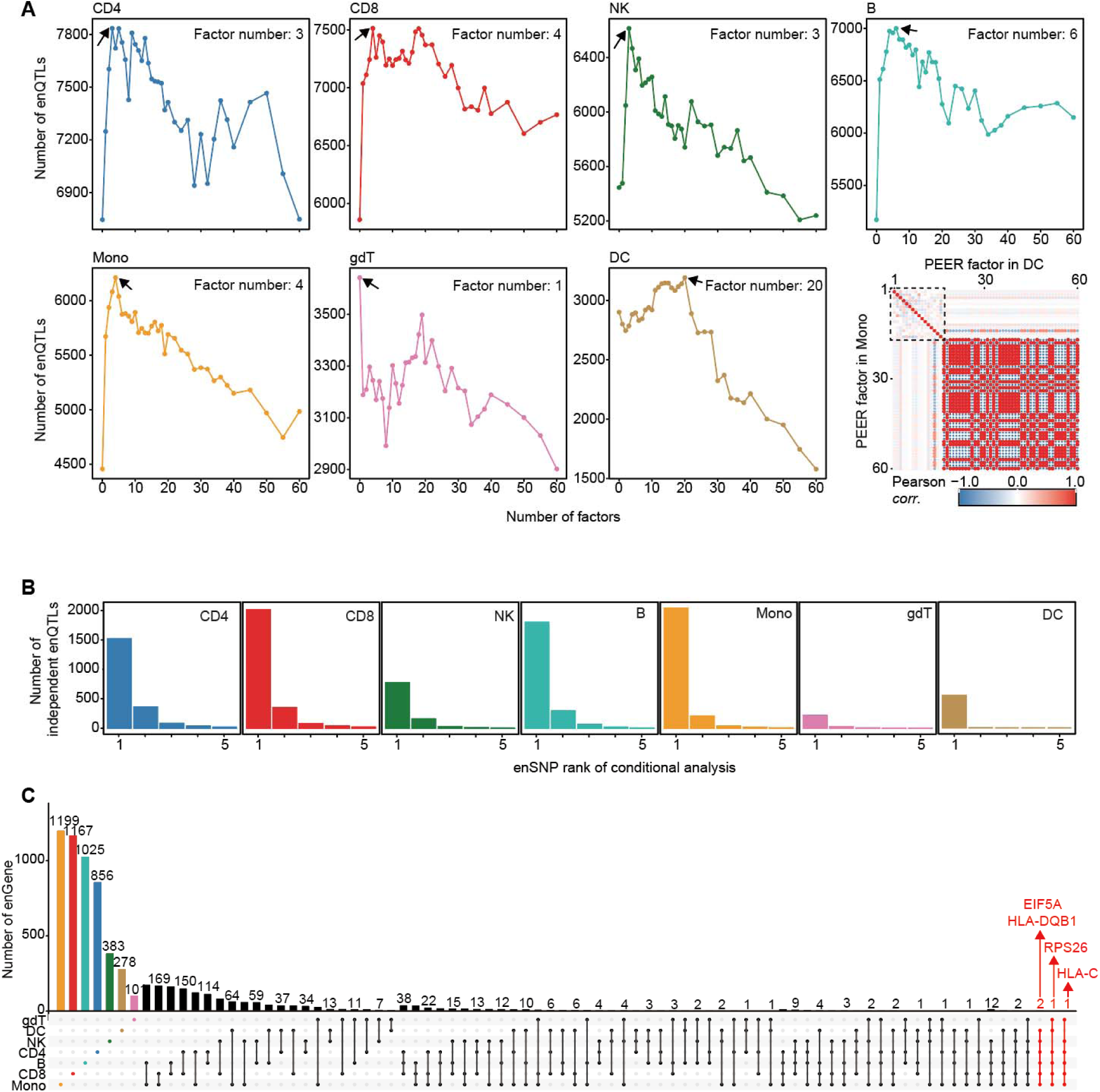
Identification of enQTL with cell-type specific PEER selection. **(A)** The number of significant enQTLs (*FDR* < 0.001) identified (y-axis) versus the number of PEER factors used (x-axis) shows the inflection point for each cell type. The graph in the lower right corner displays the Pearson correlation (color) for PEER factors in DC. **(B)** The number of independent enQTL in each round (five rounds in total) of conditional enQTL analysis are shown. **(C)** Sharing of enGenes across cell types. enGenes identified in multiple cell types are connected by lines, and the x-axis displays all categories. The red-highlighted dots represent 4 genes shared across 6 cell types.

**Figure S5.**
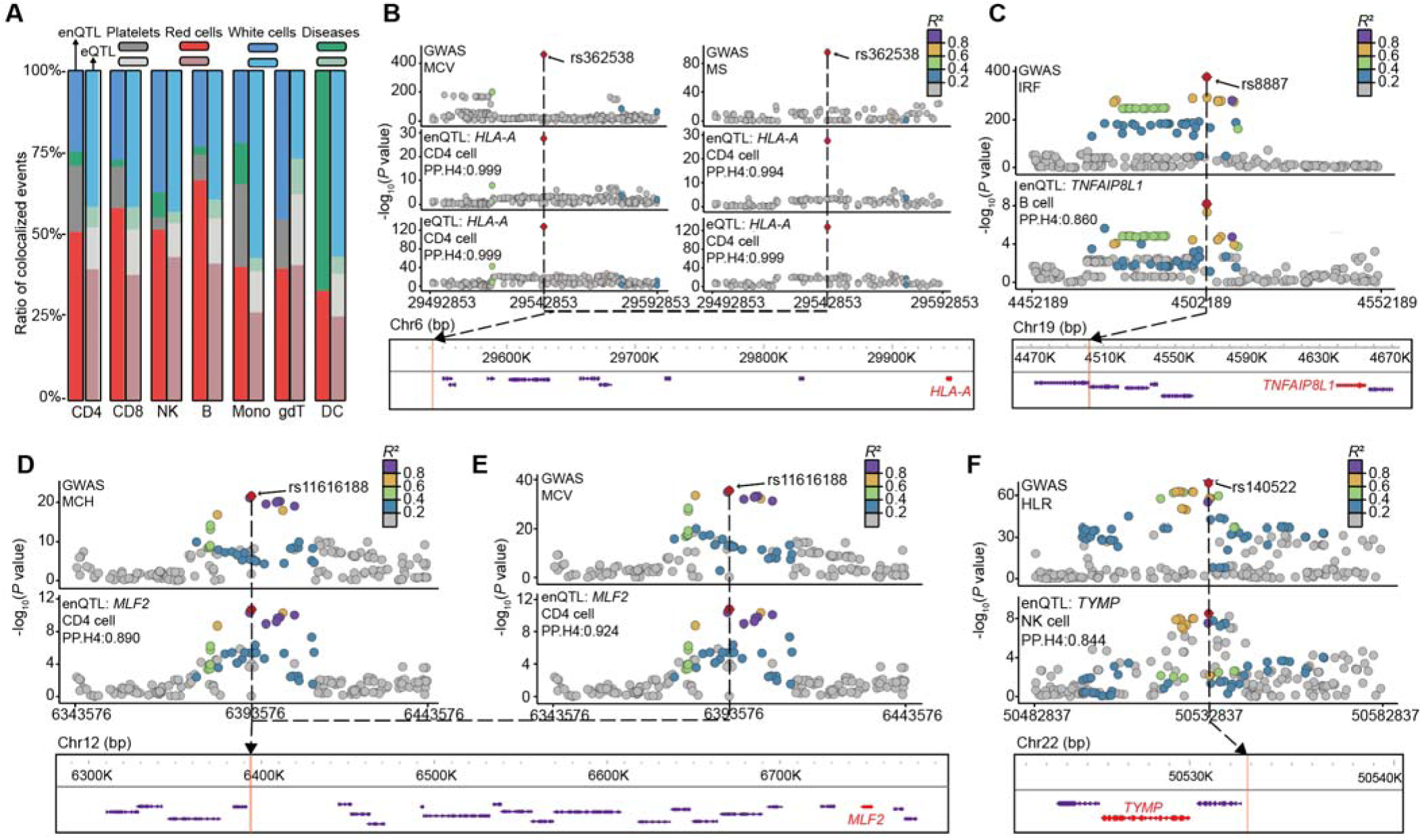
Representative loci with colocalized events between enQTL and GWAS signals. **(A)** The proportion of colocalized events across hematopoietic traits and autoimmune diseases between enQTL (dark) and eQTL (light) in different cell types. **(B)** The regional association plots of GWAS (MCV/MS), and enQTL/eQTL of the gene *HLA-A* within ±50 kb of rs362538 is presented. **(C)** The regional association plots of GWAS (IRF) and enQTL of the gene *TNFAIP8L1* within ±50 kb of rs8887 is presented. **(D-E)** The regional association plots of GWAS (MCH/MCV) and enQTL of the gene *MLF2* within ± 50 kb of rs11616188 is presented. **(F)** The regional association plots of GWAS (HLR), and enQTL of the gene *TYMP* within ±50 kb of rs140522 is presented. **Notes:** Mean corpuscular volume, MCV; Multiple sclerosis, MS; High light scatter reticulocyte count, HLR; Immature fraction of reticulocytes, IRF; Mean corpuscular hemoglobin, MCH; Mean corpuscular hemoglobin concentration.

